# Deuterium metabolic imaging and hyperpolarized ^13^C-MRI of the normal human brain at clinical field strength reveals differential cerebral metabolism

**DOI:** 10.1101/2022.02.07.22269533

**Authors:** Joshua D Kaggie, Alixander S Khan, Tomasz Matys, Rolf F Schulte, Matthew J Locke, Ashley Grimmer, Amy Frary, Martin J Graves, Mary A McLean, Ferdia A Gallagher

## Abstract

Deuterium metabolic imaging (DMI) and hyperpolarized ^13^C-pyruvate MRI (^13^C-HPMRI) are two emerging methods for non-invasive and non-ionizing imaging of tissue metabolism. Imaging cerebral metabolism has potential applications for cancer, neurodegeneration, multiple sclerosis, traumatic brain injury, stroke, and inborn errors of metabolism. Here we directly compare these two non-invasive methods at 3 T for the first time in humans, and how they simultaneously probe both glycolytic and oxidative metabolism. DMI was undertaken 1-2 hours after oral administration of [6,6’-^2^H_2_]glucose, and ^13^C-MRI was performed immediately following intravenous injection of hyperpolarized [1-^13^C]pyruvate in ten and nine normal volunteers within each arm. DMI provided maps of deuterium-labelled water, glucose, lactate, and glutamate/glutamine. ^13^C-HPMRI generated maps of hyperpolarized carbon-13 labelled pyruvate, lactate, and bicarbonate. There was clear spectral separation in the spectroscopic imaging data with both DMI and ^13^C-HPMRI at 3 T. The ratio of ^13^C-lactate/^13^C-bicarbonate (mean = 3.7 ± 1.2) acquired with ^13^C-HPMRI was higher than the equivalent ^2^H-lactate/^2^H-Glx ratio (mean = 0.18 ± 0.09) acquired with DMI. These differences can be explained by the route of administering each probe, the timing of imaging after ingestion or injection, as well as the biological differences in cerebral uptake and cellular physiology between the two molecules. The results demonstrate these two metabolic imaging methods provide different yet complementary readouts of oxidative and glycolytic metabolism within a clinically feasible timescale. Furthermore, as DMI was undertaken at a clinical field strength within a ten-minute scan time, it demonstrates its potential as a routine clinical tool in the future.

## 1. Introduction

Non-invasive methods to quantify and image cellular metabolism *in vivo* have great potential for studying the biology of many disease processes including cancer, inflammation, infection, degenerative disease, endocrine disorders, and inborn errors of metabolism. Many of these pathological processes have characteristic metabolic signatures such as aerobic glycolysis (or the Warburg effect) present in many tumors and resulting in high levels of lactate production, or the mitochondrial dysregulation and reprogramming that is present in Alzheimer’s disease (Demetrius et al., 2015; Liberti and Locasale, 2016). Positron emission tomography using the fluorine-18 labelled glucose analog fluorodeoxyglucose (^18^F-FDG-PET) is routinely used in the clinic for cancer detection and treatment monitoring, but is unable to identify the compartmentalization of the radiolabelled metabolites or the downstream products of glucose metabolism such as lactate or other metabolites (Kung et al., 2019).

Deuterium (^2^H) metabolic imaging (DMI) and hyperpolarized carbon-13 metabolic MRI (^13^C-HPMRI) are emerging techniques to probe dynamic changes in tissue metabolism using MRI without the use of ionizing radiation (Ardenkjær-Larsen et al., 2003; De Feyter et al., 2018; Vaeggemose et al., 2021; Woitek and Gallagher, 2021; Zaccagna et al., 2018). Due to the distinct biological and nuclear properties of these isotopes of hydrogen and carbon, metabolic pathways can be differentially probed using the two techniques. ^13^C has a nuclear spin of ½ and a long spin-lattice (T_1_) relaxation time of the order of many seconds *in vivo* (Daniels et al., 2016) which allows it to be pre-polarized (or hyperpolarized) before intravenous injection, allowing rapid metabolic images to be acquired dynamically over the seconds to minutes during which the substrate is delivered to the organ of interest and metabolized. In contrast, ^2^H has a quadrupolar nuclear spin of 1 and a T_1_ relaxation time of the order of hundreds of milliseconds (Ewy et al., 1986), which allows signal averaging to be performed and can be used to image slower metabolic processes (>60 min) and steady-state levels of metabolism after oral ingestion of ^2^H-labelled molecules such as glucose. However, a direct comparison between DMI and ^13^C-HPMRI in humans has not yet been undertaken, which is important for understanding the differences between these techniques and their utility for measuring glycolysis *in vivo*.

DMI is most frequently undertaken with oral deuterated [6,6’-^2^H_2_]glucose to detect the subsequent formation of its downstream metabolites: ^2^H-lactate (predominately [3,3’-^2^H_2_]lactate but also [3-H_2_]lactate), ^2^H-Glx (a combined peak of [4,4’-^2^H_2_]glutamate, [4-^2^H]glutamate, [4’-^2^H]glutamate, [4,4’-^2^H_2_]glutamine, [4-^2^H]glutamine, and [4’-^2^H]glutamine), and deuterated water (^2^H-water or HDO). *In vivo* DMI studies published to date have utilized high field strength systems greater than 3 T (De Feyter et al., 2018; de Graaf et al., 2020; Ruhm et al., 2021), but for the technique to be used more widely, translation to clinical field strengths and clinical MR systems is required. Here we demonstrate the potential of DMI at 3 T for the first time.

Hyperpolarized ^13^C-MRI is an alternative emerging method for probing tissue metabolism and utilizes dynamic nuclear polarization to increase the signal-to-noise for detection of carbon-13 labelled pyruvate by more than four orders of magnitude (Ardenkjær-Larsen et al., 2003). Hyperpolarized [1-^13^C]pyruvate is intravenously injected and tissue metabolism is non-invasively detected through the formation of downstream metabolites, such as [1-^13^C]lactate and [^13^C]bicarbonate, as measures of glycolytic and oxidative metabolism respectively. ^13^C-HPMRI can be used to image the rapid conversion of ^13^C-pyruvate within seconds, and has been used to assess normal cerebral and brain tumor metabolism in humans (Autry et al., 2020; Grist et al., 2020; Miloushev et al., 2018). The technique has many potential neurological applications such as in traumatic brain injury (DeVience et al., 2017), stroke (Xu et al., 2017), diabetes (Choi et al., 2019), and multiple sclerosis (Guglielmetti et al., 2017).

Importantly, DMI and ^13^C-HPMRI probe parallel aspects of glucose metabolism which have the potential to provide complementary biological information in the brain (**Figure 1**). Labelled ^2^H-glucose is rapidly transported across the blood brain barrier (BBB) by GLUT1, an isoform of the glucose transporter, whereas ^13^C-pyruvate is transported across the BBB predominately by the monocarboxylic acid transporter, MCT1 (Bergersen et al., 2002; Duelli and Kuschinsky, 2001). Each glucose molecule undergoes glycolysis to two molecules of pyruvate and the ^13^C-pyruvate or ^2^H-pyruvate exchanges with ^13^C-lactate or ^2^H-lactate in the cytosol, catalyzed by the enzyme lactate dehydrogenase (LDH). Alternatively, pyruvate may enter the mitochondrion undergoing oxidative metabolism where the two labelled pathways diverge: the ^13^C label irreversibly forming ^13^C-labelled carbon dioxide catalyzed by pyruvate dehydrogenase (PDH) and detected through the subsequent formation of ^13^C-bicarbonate, while the ^2^H-label passes into the tricarboxylic acid (TCA) cycle forming α-ketoglutarate (^2^H-αKG) and downstream ^2^H-glutamate and/or ^2^H-glutamine, detected as a combined ^2^H-Glx signal (Melkonian and Schury, 2019). Comparisons between the two techniques have been undertaken in a murine model, showing an inverse relationship between ^13^C-lactate and ^2^H-Glx (von Morze et al., 2021). Here we have undertaken a comparison of DMI and ^13^C-HPMRI in humans for the first time by applying them to the healthy human brain at 3 T to explore the differences between the two imaging methods.

**Figure 1.**
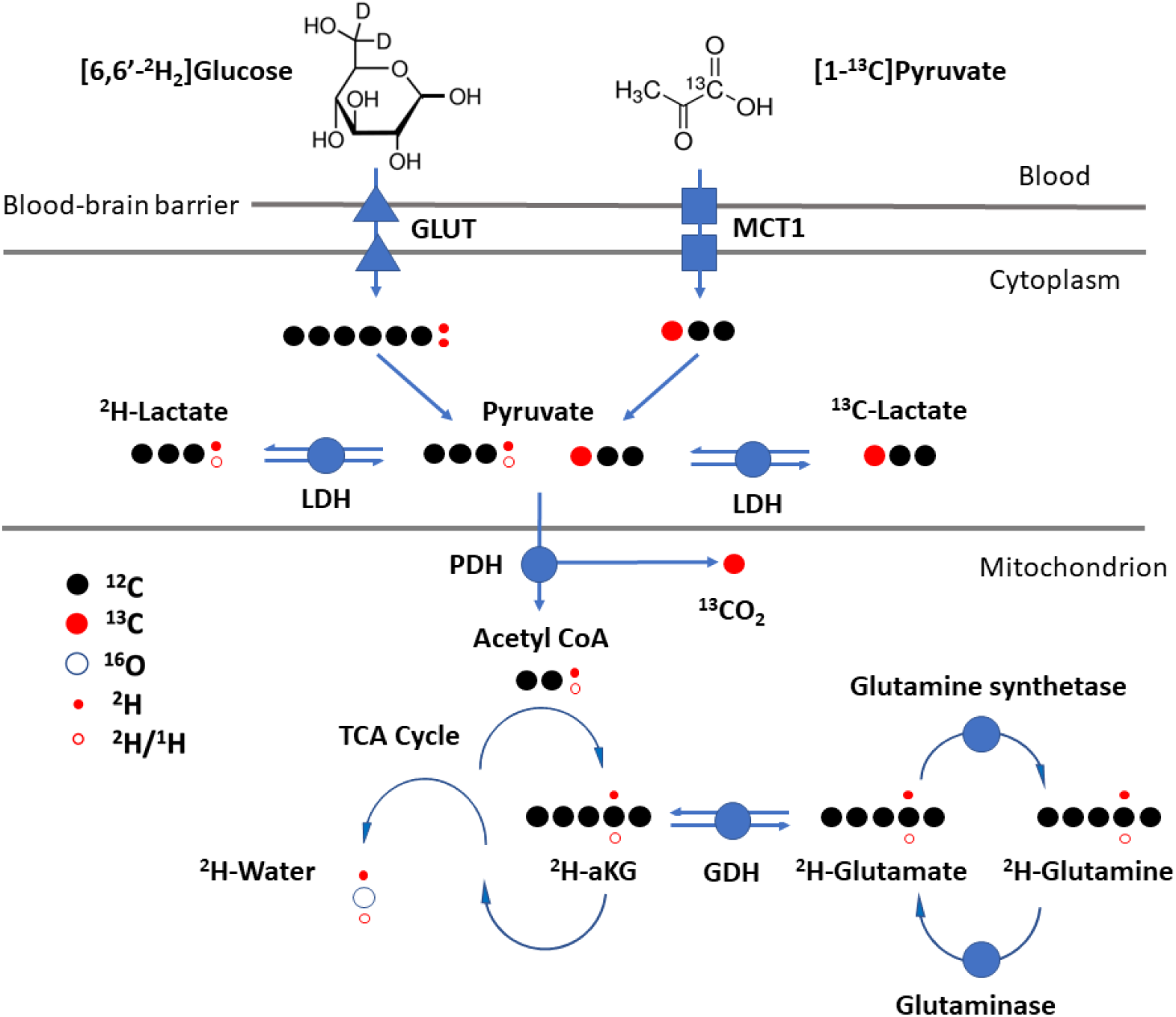
Schematic showing the differential metabolic pathways of [6,6’-^2^H_2_]glucose metabolism detected with DMI, and [1-^13^C]pyruvate metabolism detected with ^13^C-HPMRI. GLUT1 and MCT1 are the main transporters of glucose and pyruvate across the blood brain barrier respectively. Glycolysis is undertaken in the cytosol, converting glucose into pyruvate. Lactate dehydrogenase (LDH) and pyruvate dehydrogenase (PDH) catalyze the conversion of pyruvate into either lactate or acetyl coenzyme A (Acetyl CoA) and carbon dioxide respectively. Glutamate dehydrogenase (GDH) catalyzes the conversion of the tricarboxylic acid (TCA) intermediate α-ketoglutarate (αKG) into Glx. Black circles represent carbon nuclei, red circles either labelled ^13^C nuclei or ^2^H deuterons, and small open circles represent protons or deuterons reflecting the different combinations of labelled molecules that may be present.

## 2. Material and Methods

Hyperpolarized ^13^C-MRI and ^2^H-glucose DMI were performed using a 3 T MRI system (MR750; GE Healthcare, Waukesha, WI, USA). Eleven normal volunteers in total were imaged (male:female 7:4; age 31±7 years), with ten undergoing DMI and nine undergoing ^13^C-HPMRI. Eight subjects overlapped between groups undergoing both DMI and ^13^C-HPMRI. The study was performed with the approval of a local research ethics committee (Molecular Imaging and Spectroscopy with Stable Isotopes in Oncology and Neurology: MISSION-MIMS; REC: 15/EE/0255) and with the informed consent of all volunteers.

All starting materials for both the DMI and ^13^C-HPMRI probes were obtained at the pharmaceutical grade required for oral consumption and intravenous injection respectively. For the DMI, [6,6’-^2^H_2_]glucose solution was prepared immediately before oral consumption by the participant. For the ^13^C-HPMRI, pharmacy kits containing [1-^13^C]pyruvic acid were assembled in aseptic conditions using validated processes in a cleanroom as described previously (Grist et al., 2019). Additional safety checks, including pH measurements, were performed before injection of the ^13^C-labelled pyruvate. Blood glucose was measured using a finger-prick blood sample before and after consumption of the ^2^H-labelled glucose.

Prior to imaging each X-nucleus, T_1_W 3D proton (^1^H) images using a spoiled gradient recalled echo sequence were acquired and high-order shimming was performed. Eddy current settings were then adjusted for either ^13^C or ^2^H (McLean et al., 2021). Bloch-Siegert radiofrequency calibration measurements were then performed for both ^13^C and ^2^H to determine the transmit power and imaging frequencies (Sacolick et al., 2010).

### 2.1 Deuterium Metabolic Imaging (DMI)

3D density-weighted magnetic resonance spectroscopic imaging (MRSI) and unlocalized magnetic resonance spectroscopy (MRS) were used to obtain metabolite concentration estimates and mapping of water (4.7 ppm), glucose (3.9 ppm), Glx (2.4 ppm), lactate (1.35 ppm), and lipids (0.9 ppm) (Lu et al., 2017). MRS was used to obtain this data over the entire brain volume, while MRSI acquired localized spectroscopic data with 3.2 cm isotropic voxels. The DMI study arm used a four-rung ^2^H birdcage head coil with a 30 cm diameter and 24 cm rung length. The birdcage and hybrid 0-90° power splitter were developed in-house, and combined with a commercial transmit/receive switch (Clinical MR Solutions, Brookfield, Wisconsin, USA).

Human experiments were performed after the oral ingestion of [6,6’-^2^H_2_]glucose (^2^H-glucose, Cambridge Isotopes Laboratories, Inc., Tewksbury, USA) following a minimum of 6 hours of fasting to ensure rapid uptake of the glucose bolus. The ^2^H-glucose was administered orally based on weight at a dose of 0.75 g/kg up to a maximum of 60 g per volunteer, and dissolved in 200 mL of potable water. A single dynamic experiment was performed with one subject at baseline and at 12 timepoints to repeatedly measure the deuterium uptake immediately after oral ingestion to determine an optimal imaging time point for deuterium. The first water measurement was assumed to have a deuterated water concentration of 10.9 mM based on previous publications (Ashwal et al., 2004). The data obtained from the first DMI subject at 57 min post-ingestion were included with data from all subjects for comparative measurements.

Ten subjects were imaged 57-136 min following oral ^2^H-glucose administration using 3D MRSI with the following parameters: field-of-view (FOV) = 32 cm, TR = 120 ms, flip angle = 90°, 5000 Hz, 544 points, matrix = 10×10×10 with 2 averages of 2504 transients, weighted with a Hamming filter. Unlocalized MR spectra were also acquired prior to and following the MRSI with the following parameters: flip = 90°, TR=1 s, 128 averages, 5000 Hz, 2048 points.

### 2.2 Hyperpolarized ^13^C-MRI

The normal volunteers in the ^13^C-HPMRI arm of the project were injected with an intravenous bolus of [1-^13^C]pyruvate: 0.4 mL/kg of ∼250 mM pyruvate, up to 40 mL at rate of 5 mL/s as described previously (Grist et al., 2019). The ^13^C-pyruvic acid was hyperpolarized using a SpinLab hyperpolarizer (Research Circle Technology, Albany NY, USA). ^13^C-MRI was performed using a dual-tuned 16-rung ^13^C/^1^H quadrature transmit/receive head coil (Rapid Biomedical, Rimpar, Germany), with a 30 cm diameter and 24 cm rung length.

2D MRSI acquisition was performed with the following parameters: FoV = 20 cm, TR = 323 ms, flip angle = 8-10°, matrix = 10×10 with 61 transients, 5 slices, 2 cm thick, commencing between 22-27 s following injection. Unlocalized spectra were acquired following the MRSI acquisition: TR = 1 s, flip angle = 45°, 5000 Hz, 2048 points, 1 to 8 transients collected beginning 44-49 s after pyruvate injection.

### 2.3 Data processing

MRS and MRSI data were analyzed in MATLAB using the OXSA AMARES toolbox (Purvis et al., 2017) to fit spectral peak areas and create metabolite maps, with a signal-to-noise (SNR) threshold of 2 for voxel rejection. ^2^H-glucose, ^2^H-lactate, and ^2^H-Glx images were normalized to the maximum water signal over the entire image and as ratio maps to the local water signal. The water ratio maps accounted for differences in signal non-uniformities for quantitative comparisons. ^13^C-pyruvate, ^13^C-lactate, and ^13^C-bicarbonate metabolite maps were normalized to the peak pyruvate value and whole-brain values were normalized to the total ^13^C signal (summed signals from pyruvate, lactate and bicarbonate). Interpolation of the deuterium MRSI was performed using zero-filling to generate the same voxel size as acquired with the ^13^C-MRSI. Derived metabolite images were upscaled via zero-filling from 10×10 to 16×16 and were also smoothed using a median filter for display.

Pearson linear correlations and significance were calculated between the changes of metabolites over time in the first subject. Paired t-tests were used to compare whether there were significant differences between the ^2^H-lactate/Glx values measured with whole brain MRS and those averaged from the MRSI voxels within the brain. Paired t-tests were also used to compare blood glucose drawn prior to and immediately following imaging, and to compare ^13^C-lactate/bicarbonate with ^2^H-lactate/Glx.

## 3. Results

### 3.1 Timecourse of cerebral DMI measurements

Dynamic DMI measurements were undertaken in a single volunteer following glucose administration to assess the optimal time for subsequent data acquisition. Deuterated water, glucose, Glx, and lactate signals had positive, significant linear correlations with time (*r*^*2*^ ≥ 0.70, *p*-values ≤ 0.002) over the first 60 min after oral intake (**Figure 2**). ^2^H-glucose increased by 8x up to a maximum of 7.8 mM, referenced against an assumed baseline HDO signal of 10.9 mM (Ashwal et al., 2004). This was accompanied by a smaller rise by 6x in ^2^H-Glx (up to 3.0 mM). HDO increased slightly by <12% (up to 11.2 mM). A small peak at 1.3 ppm was observed to increase by 71% relative to the timepoint at 15 min (up to 1.3 mM), or by 91% relative to baseline, and was attributed to a combination of the lactate and lipid signal. The lipid signal at 0.9 ppm did not show a significant increase (*r*^*2*^ = 0.39, *p*-value = 0.17). The rate of increase in glucose concentration appeared to decrease by the end of the hour, suggesting that it was starting to plateau.

**Figure 2.**
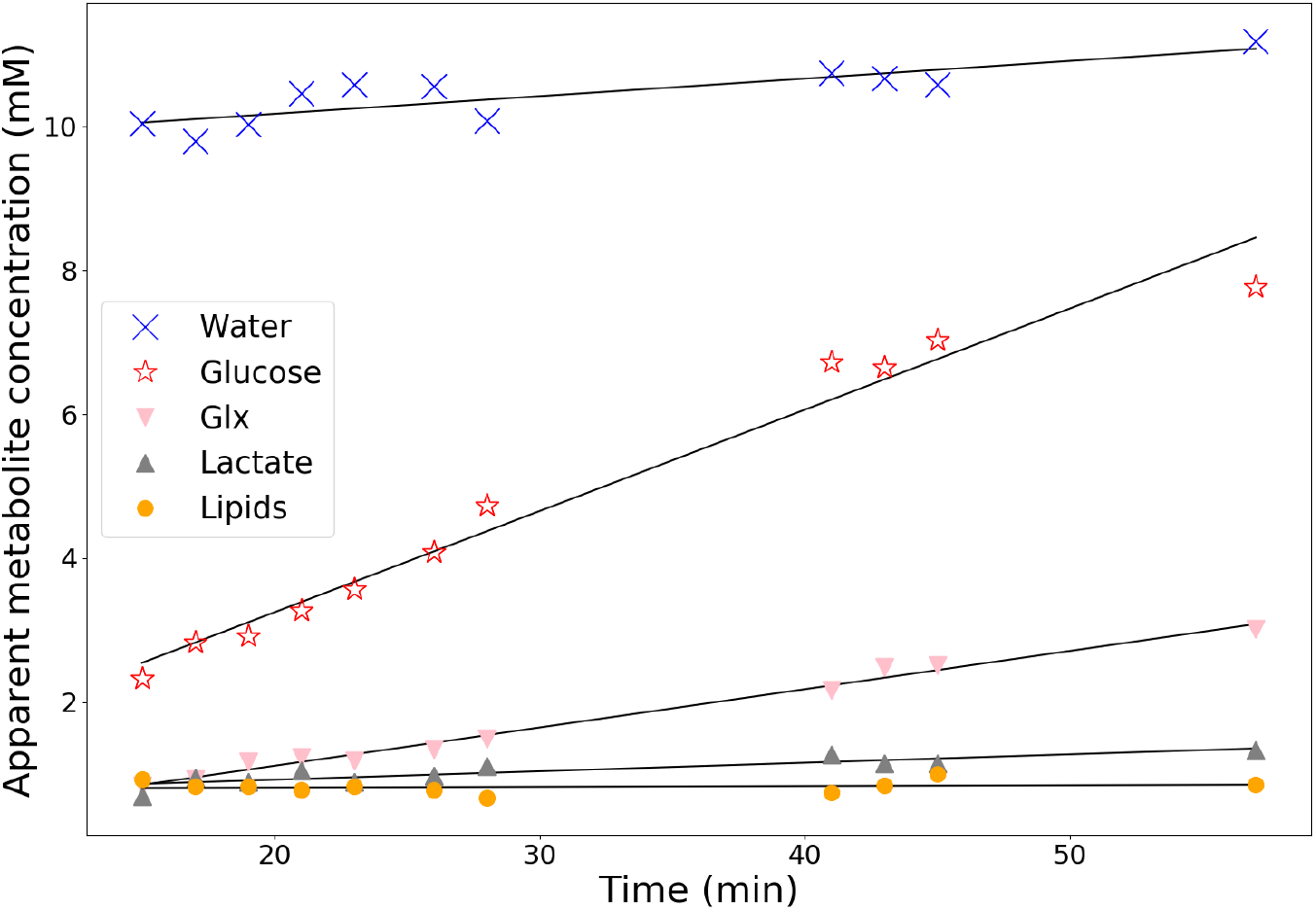
Timecourse of the concentration of deuterated glucose and its downstream metabolites over the first hour following oral ingestion in the brain of a single volunteer (HV1). The baseline signal from labelled water (HDO) was assumed to be 10.9 mM, and other metabolites were referenced against this as no additional calibrations were performed between scans. Glx: combined signal from glutamine and glutamate. Linear regression models: [^2^H-glucose] = 0.14t+0.43, p-value < 10^−7^; [^2^H-Glx] = 0.053t+0.049, p-value < 10^−9^; [^2^H-lactate] = 0.012t+0.69, p-value < 10^−3^; and [^2^H-lipids] = 0.001t+0.792, p-value = 0.17.

Based on these initial results, the remaining subjects were imaged at least 1 hour after the administration of glucose to maximize signal from the downstream metabolites, while minimizing the time required for the volunteers to remain in the magnet. Overall DMI measurements were obtained 107 ± 21 min after ingestion of glucose (**Table 1**), which enabled changes in metabolism over the course of the second hour to be evaluated.

**Table 1.** Metabolite ratios across all subjects measured using unlocalized ^2^H-MRS acquisition derived by summing the spectra from 5 slices and processed using AMARES.

| Subject | Age Range (years) | Gender | Volunteer weight (kg) | Time after consumption (min) | Baseline blood glucose (mmol/L) | Post blood glucose (mmol/L) | Glucose/HDO | Glx/HDO | Lactate/HDO | Lactate/Glx | HDO linewidth (Hz) |
| --- | --- | --- | --- | --- | --- | --- | --- | --- | --- | --- | --- |
| HV1 | 36-40 | F | 67 | 57 | 4.7 | 7.8 | 0.69 | 0.27 | 0.12 | 0.44 | 13.8 |
| HV2 | 18-25 | M | 80 | 114 | 5.7 | - | 0.68 | 0.40 | 0.06 | 0.16 | 10.5 |
| HV3 | 26-30 | M | 82 | 104 | 5.8 | - | 0.25 | 0.28 | 0.03 | 0.14 | 11.8 |
| HV4 | 41-45 | F | 59 | 120 | 5.8 | - | 0.27 | 0.30 | 0.03 | 0.12 | 11.0 |
| HV5 | 18-25 | M | 81 | 94 | 4.3 | 7.6 | 0.42 | 0.20 | 0.05 | 0.25 | 13.8 |
| HV6 | 26-30 | F | 77 | 121 | 5.6 | 6.7 | 0.33 | 0.23 | 0.02 | 0.12 | 11.5 |
| HV7 | 31-35 | M | 82 | 136 | 5.9 | 6.2 | 0.70 | 0.35 | 0.07 | 0.18 | 12.7 |
| HV8 | 26-30 | M | 65 | 115 | 6.8 | - | 0.39 | 0.28 | 0.05 | 0.18 | 12.4 |
| HV9 | 31-35 | M | 101 | 130 | 5.9 | 4.6 | 0.23 | 0.25 | 0.03 | 0.12 | 13.1 |
| HV10 | 26-30 | F | 58 | 105 | 5.2 | 4.9 | 0.31 | 0.24 | 0.02 | 0.10 | 11.7 |
| **Mean** | 31±7 | | 75±12 | 109±21 | 5.6±0.7 | 6.3±1.2 | 0.43±0.18 | 0.28±0.06 | 0.05±0.03 | 0.18±0.09 | 12.3±1.2 |

### 3.2 DMI cerebral metabolism measurements

Signal from both ^2^H-lactate and ^2^H-Glx was detected in all volunteers >57 min after oral administration of labelled glucose. ^2^H-lactate was used as a measure of cytosolic glycolytic metabolism and ^2^H-Glx as a measure of mitochondrial oxidative metabolism.

An unlocalized whole-brain ^2^H-spectrum from a single subject is shown in **Figure 3A** alongside that acquired using ^2^H-MRSI from a single voxel (3.2×3.2×3.2 cm^3^) near the center of the brain for comparison, with the location indicated on the proton image (**Figure 3B**). The ^2^H signals from the individual metabolites in the whole-brain data are subject to line broadening from magnetic non-uniformities across the brain. In contrast, separate peaks from HDO, ^2^H-glucose, ^2^H-Glx, and ^2^H-lactate can be more easily resolved in the single voxel spectrum obtained using MRSI. The ^2^H-lactate+lipid peak at 1.3 ppm was small in all the volunteers but could be detected above the noise floor, and was larger than the 0.9 ppm ^2^H-lipid peak, as reported previously (De Feyter et al., 2018). Additionally, the ^2^H-glucose/^2^H-water ratio in the global spectrum was approximately 10% higher than in the MRSI midline voxel, secondary to the effects of the broader unlocalized spectra.

**Figure 3.**
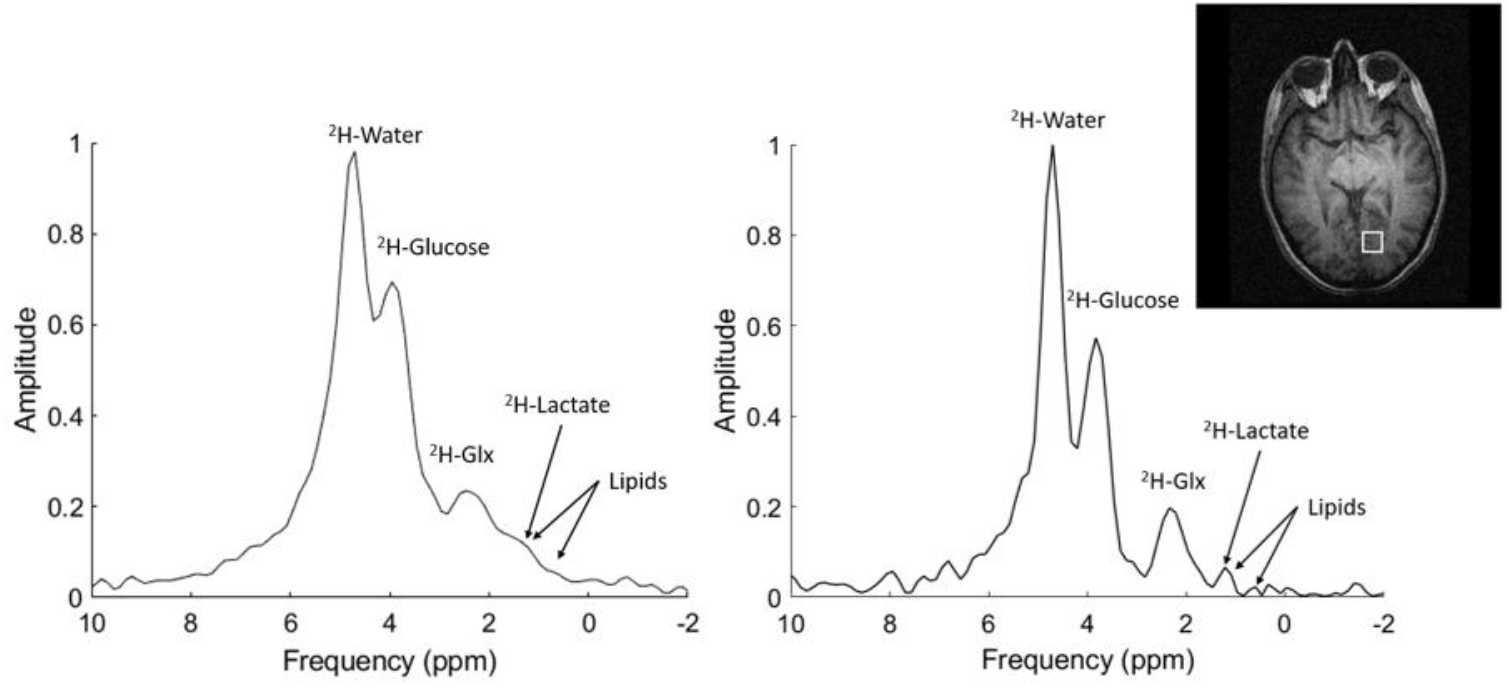
DMI spectra acquired 90 min after oral administration of ^2^H-glucose in a normal volunteer using 3 T MRI (HV2). Left: Whole-brain ^2^H-spectra. Right: Localized ^2^H-spectra acquired from a single midline voxel (3.2 cm isotropic). The localized spectrum demonstrates that ^2^H-water (4.7 ppm), ^2^H-glucose (3.9 ppm), ^2^H-Glx (2.4 ppm), and ^2^H-lactate (1.35 ppm) peaks were more easily detected than on the global spectrum because of line broadening from magnetic non-uniformities causing. The lipids peak at 0.9 ppm was close to the noise floor.

The individual volunteer data acquired from whole-brain spectroscopy are shown in **Table 1**. The ratio of oxidative to glycolytic metabolism in the whole-brain spectra favored oxidation in all volunteers, with a high level of ^2^H-Glx detected across the brain (mean ± standard deviation of the ^2^H-lactate/^2^H-Glx ratio = 0.19 ± 0.12); similar results were obtained when the MRSI metabolite maps were used to obtain this ratio (mean MRSI ^2^H-lactate/^2^H-Glx ratio = 0.24 ± 0.05, p = 0.08). There was an approximate linear correlation between the ^2^H-lactate/^2^H-Glx ratio and time after ingestion, which demonstrates that mitochondrial metabolism takes place on a slower timescale compared to glycolytic metabolism in the cytosol (r^2^ = 0.69, p = 0.002, Supplementary **Figure S1**).

Deuterium-labelled water, glucose, Glx, and lactate maps derived from the ^***2***^H-MRSI spectra acquired following the administration of ^2^H-glucose are shown in **Figure 4**. The ^2^H signal was slightly asymmetrical across the brain, with increased signal of all metabolites on the right side of the image. As B_1_ mapping was not performed, accurate quantitation is not possible over the entire volume. For quantitation, the maps of each subject were referenced with respect to HDO to obtain relative metabolite maps. **Figure 4** shows ratiometric maps with relative metabolic homogeneity across the brain.

**Figure 4.**
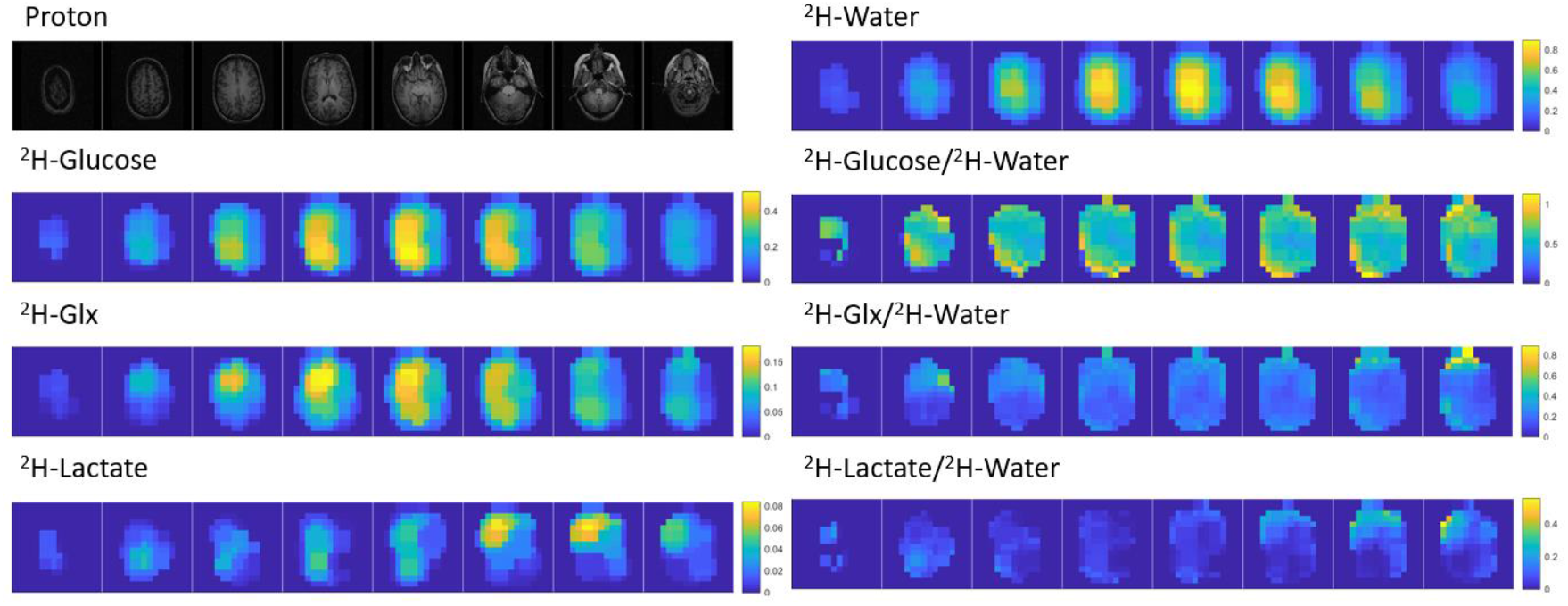
3D T_1_-weighted proton (^1^H) images and metabolite maps acquired 105 mins after administration of ^2^H-glucose in volunteer HV4. Metabolite levels were normalized to the ^2^H-water (HDO) signal. Images were upscaled using zero filling to generate 2.0×2.0×3.2 cm^3^ voxels for peak fitting, then metabolite maps were smoothed with a median filter. The metabolite maps on the left were scaled to the maximum water signal detected in the brain. Due to coil sensitivity differences, metabolite ratios to water were used for quantitation.

There were no significant differences in blood glucose measured prior to and immediately following imaging (5.6 ± 0.7 mmol/L baseline, 6.6 ± 1.1 mmol/L post-imaging, p = 0.23).

### 3.3 Measurements of cerebral metabolism with ^13^C-HPMRI

^13^C-pyruvate signal, and its downstream metabolites ^13^C-lactate and ^13^C-bicarbonate, were detected using MRSI in the brains of all volunteers after intravenous injection of hyperpolarized [1-^13^C]pyruvate. An unlocalized whole-brain ^13^C-spectrum from a single subject is shown in **Figure 5A** alongside the spectrum from a single voxel, with the voxel location indicated on the proton image (**Figure 5B**). Individual volunteer data obtained using ^13^C-HPMRI are shown in **Table 2**. ^13^C-lactate signal was used as a measure of cytosolic glycolytic metabolism and ^13^C-bicarbonate as a measure of mitochondrial oxidative metabolism. In contrast to DMI, the ratio of glycolytic to oxidative metabolism using ^13^C-HPMRI favored the former, with a mean (± standard deviation) ^13^C-lactate/^13^C-bicarbonate ratio of 3.7 ± 1.3.

**Figure 5.**
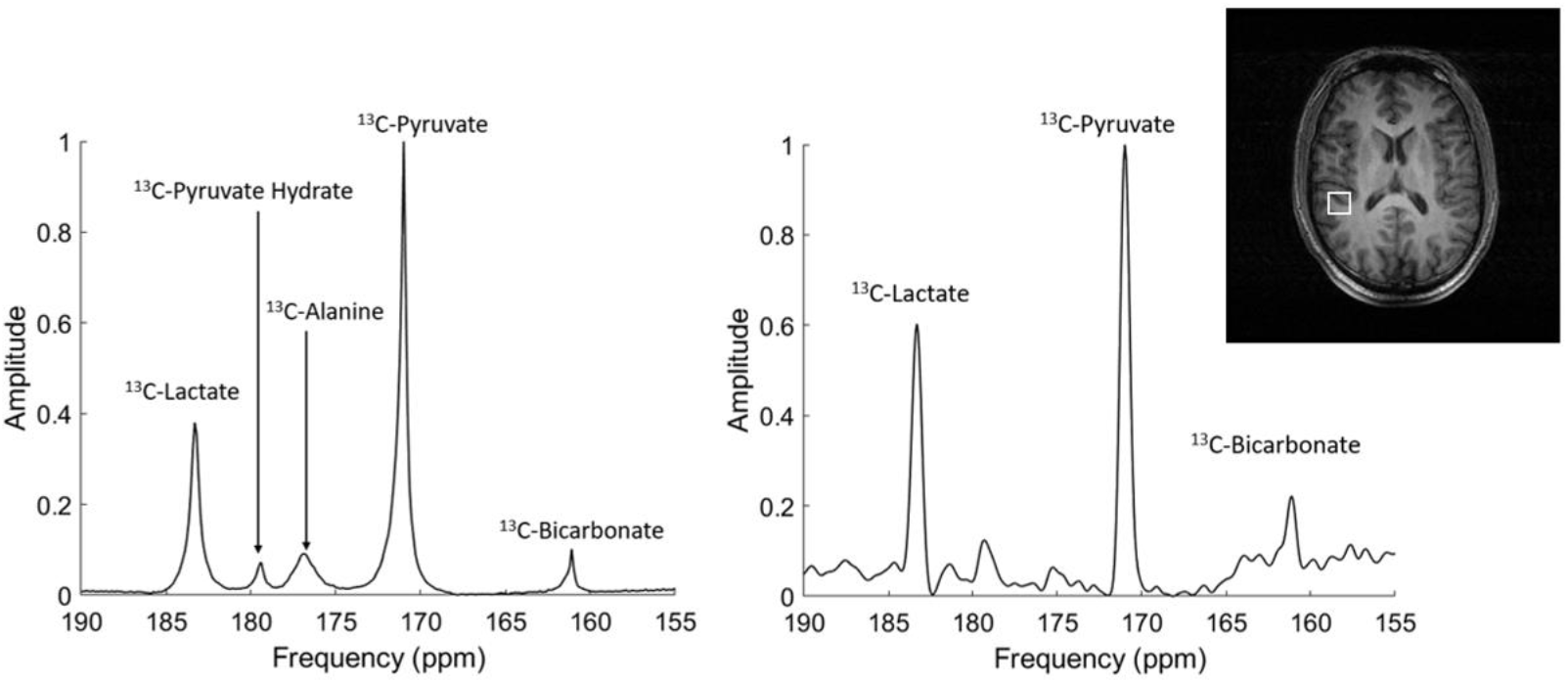
^13^C-HPMRI spectra after intravenous administration of ^13^C-pyruvate acquired in a normal volunteer (HV7) using 3 T MRI. Left: Whole-brain ^13^C-spectra 49 s after injection. Right: Localized ^13^C-spectra acquired from a single MRSI voxel (2 cm isotropic). The spectra demonstrate clear differentiation of ^13^C-pyruvate (171 ppm), ^13^C-lactate (183 ppm), and ^13^C-bicarbonate (161 ppm) with both MRS and MRSI. ^13^C-pyruvate hydrate (179 ppm) and ^13^C-alanine (177 ppm) are detectable in the global MRS spectrum, but alanine is not seen in the local MRSI spectrum.

**Table 2.** Metabolite ratios derived from unlocalized whole-brain spectroscopy following ^13^C-MRSI and measured 44-49 s after injection. The values reported for ^13^C-pyruvate, ^13^C-lactate, and ^13^C-bicarbonate are normalized to the total ^13^C-signal. ^13^C-HPMRI was performed in HVs 1-8 and 11, while DMI was performed in HVs 1-10.

| Volunteer number | Polarization (%) | Pyruvate | Lactate | Bicarbonate | Lactate/Pyruvate | Bicarbonate/Pyruvate | Lactate/Bicarbonate |
| --- | --- | --- | --- | --- | --- | --- | --- |
| HV1 | 12.7 | 0.60 | 0.30 | 0.10 | 0.50 | 0.17 | 3.03 |
| HV2 | 15.1 | 0.72 | 0.20 | 0.09 | 0.27 | 0.12 | 2.27 |
| HV3 | 6.1 | 0.54 | 0.38 | 0.08 | 0.70 | 0.15 | 4.76 |
| HV4 | 13.4 | 0.74 | 0.21 | 0.06 | 0.28 | 0.08 | 3.70 |
| HV5 | 22.8 | 0.54 | 0.37 | 0.10 | 0.68 | 0.18 | 3.85 |
| HV6 | 18.7 | 0.70 | 0.25 | 0.06 | 0.36 | 0.08 | 4.35 |
| HV7 | 13.4 | 0.67 | 0.27 | 0.07 | 0.40 | 0.10 | 4.00 |
| HV8 | 19.3 | 0.48 | 0.44 | 0.07 | 0.92 | 0.15 | 5.88 |
| HV11 | 6.1 | 0.76 | 0.14 | 0.11 | 0.18 | 0.14 | 1.32 |
| **Mean** | **14±5** | **0.63±0.09** | **0.28±0.09** | **0.08±0.02** | **0.48±0.23** | **0.13±0.03** | **3.68±1.21** |

The ^13^C metabolite maps derived from MRSI in a single volunteer are shown in **Figure 6** with axial images of ^13^C-pyruvate, ^13^C-lactate, and ^13^C-bicarbonate collected 27 s following intravenous injection of hyperpolarized ^13^C-pyruvate in five slices through the brain. The metabolite maps in **Figure 6** show higher signal for all three ^13^C-labelled metabolites peripherally within the brain compared to centrally. This suggests higher production of lactate and bicarbonate in gray matter compared to white, which is in agreement with previous work and may be secondary to higher perfusion and therefore pyruvate delivery in these areas (Dwork et al., 2021; Grist et al., 2019; Lee et al., 2020; Ma and Park, 2020). The signal of all three metabolites was especially high in the midline posteriorly and superiorly, which is likely to reflect venous drainage and the distribution of the cerebral venous sinuses (Grist et al. 2019).

**Figure 6.**
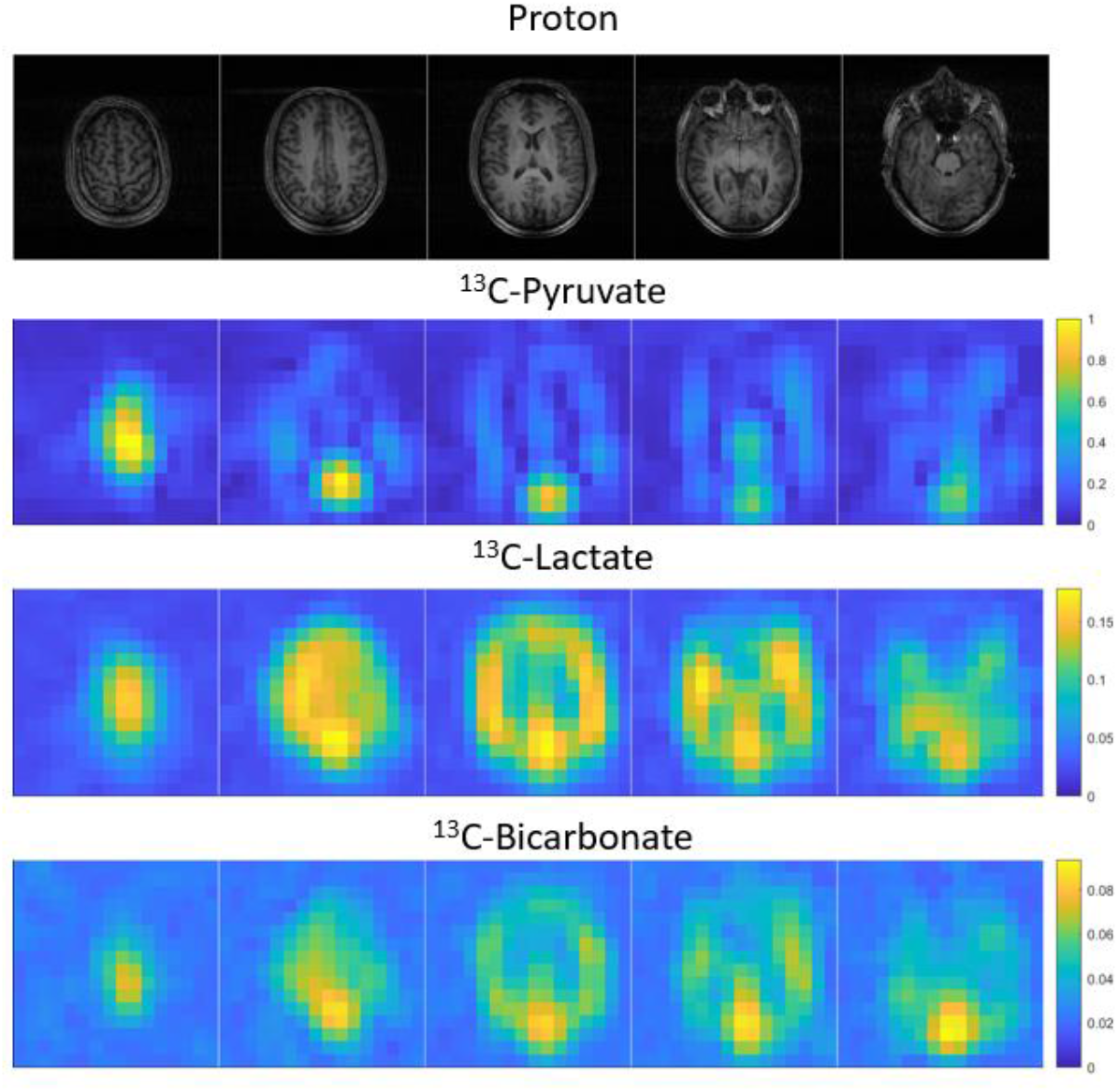
3D T1-weighted proton images (top row) and metabolite maps of the brain acquired using ^13^C-MRSI 27 s after injection of hyperpolarized ^13^C-pyruvate in HV5. The signal has been normalized to the maximum total ^13^C-signal.

### 3.4 Comparison between DMI and ^13^C-HPMRI

Acquisition of both DMI and ^13^C-HPMRI was undertaken in eight subjects. Both sets of images acquired from the same normal volunteer (HV8) are shown in **Figure 7** for direct comparison. The HDO signal was symmetrical demonstrating uniformity of both the coil sensitivity and the labelled water distribution throughout the brain, approximately two hours following labelled glucose administration. There are observable differences between HDO and ^2^H-glucose signal distribution, as demonstrated by the calculated ratio map of the two, indicating that glucose accumulation in tissue is dependent on factors in addition to delivery of the probe. This case also demonstrated even more marked differences in the distribution of ^2^H-glucose and ^2^H-lactate in keeping with differential glucose metabolism across the brain, with particularly high levels of ^2^H-lactate laterally. A comparison of the ^13^C-pyruvate and ^2^H-glucose distribution also showed differences, with the ^2^H-glucose more uniform than the ^13^C-pyruvate, with the latter more likely to be influenced by the vascular distribution. Furthermore, comparison between the ^13^C-lactate and ^2^H-lactate maps also showed differences in the distribution of the metabolite using the two techniques, with higher ^13^C-lactate signal seen centrally, compared to the more peripheral ^2^H-lactate signal. The ^2^H-glucose and ^2^H-Glx images were also more homogeneous in appearance compared to the ^13^C-lactate and ^2^H-lactate images. No significant linear correlations (p>0.05) were observed between any of the ^2^H and ^13^C metabolites in volunteers that had both imaging undertaken: the strongest trend was a negative correlation between ^13^C-lactate/bicarbonate and ^2^H-lactate/Glx (*r*^2^ = 0.14, *p*-value=0.41), and is shown in Supplementary **Figure S2**.

**Figure 7.**
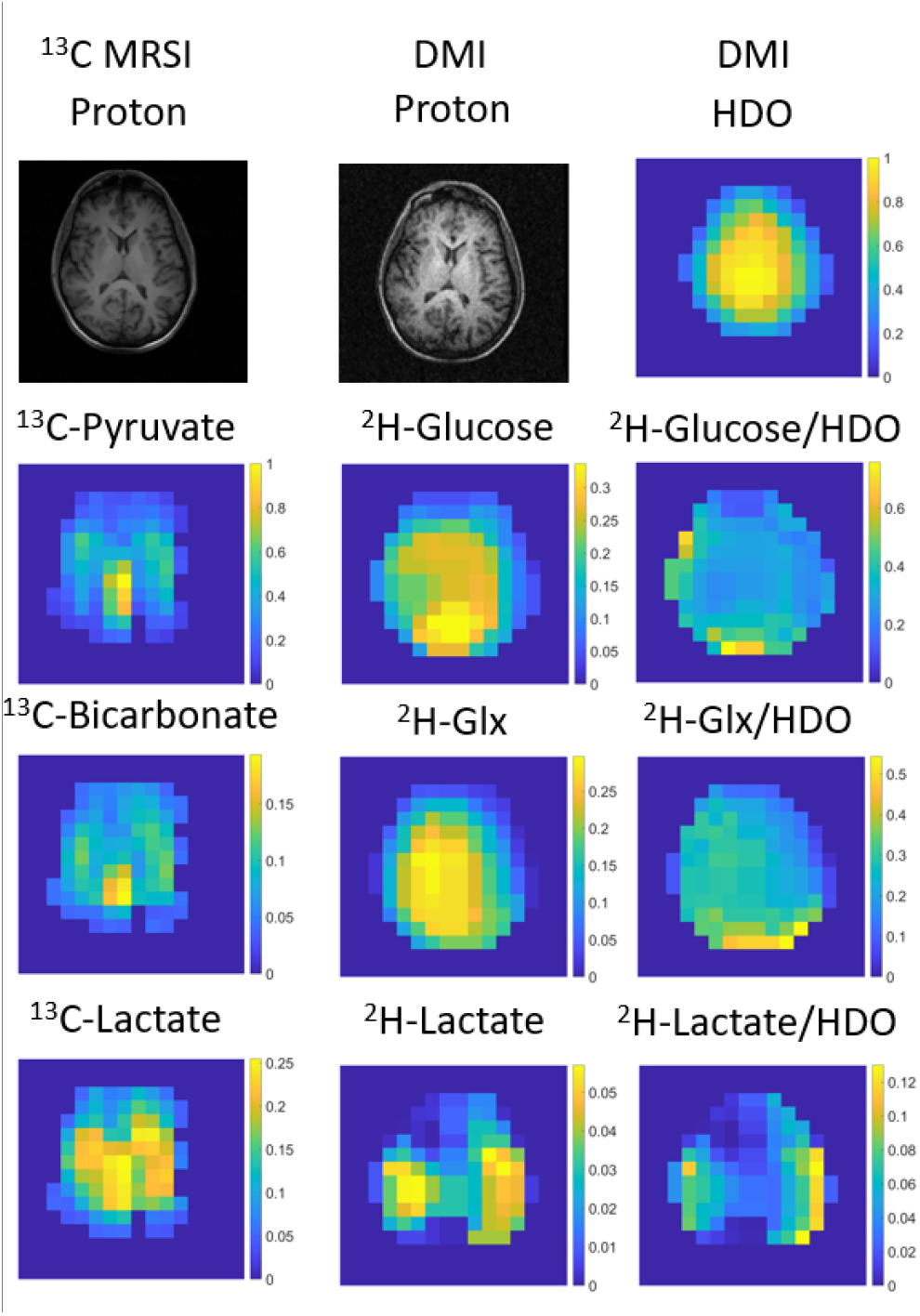
Comparison of DMI and ^13^C-HPMRI in one of the eight volunteers who underwent both imaging approaches (HV8). DMI was performed 115 min after ingestion of ^2^H-glucose. There was significant variation in the distribution of the metabolites across the brain: for example the ^2^H-glucose and ^2^H-Glx maps were more homogeneous in appearance than the ^13^C-lactate and ^2^H-lactate images.

## 4. Discussion

Although glucose is the major fuel source in the brain, both pyruvate and lactate are important intermediaries in cerebral energy metabolism. ^18^F-FDG-PET has been an important imaging tool to study the uptake and phosphorylation of glucose in the brain, but it does not provide information on downstream metabolites, or the role played by monocarboxylic acids. DMI and ^13^C-HPMRI are two emerging techniques for non-invasively probing these alternative aspects of carbohydrate metabolism which could have important implications for evaluating cerebral energy utilization in health and disease. However, the way in which these techniques differ in probing human brain metabolism has not been studied previously. One of the major challenges in undertaking a direct comparison between the two techniques in humans is that ^13^C-HPMRI is usually undertaken at 3 T, which allows good spectral separation of metabolites while maintaining a sufficiently long relaxation time for the hyperpolarized signal to enable detection of both the injected hyperpolarized substrate and the subsequently formed metabolites (Vaeggemose et al., 2021). In comparison, DMI has been applied at field strengths of up to 16.4 T for improved spectral separation and signal-to-noise (Lu et al., 2017), with clinical DMI having previously been undertaken at field strengths of 4 T and higher, which are not used in routine clinical practice (De Feyter et al., 2018; de Graaf et al., 2020; Ruhm et al., 2021).

This study has shown for the first time the feasibility of performing clinical DMI with orally administered ^2^H-glucose at 3 T. The acquired spectra could be used to resolve ^2^H-glucose, ^2^H-water, ^2^H-lactate, and ^2^H-Glx in the normal brain using spectroscopic imaging, especially within localized voxels, demonstrating its potential to reveal regional variations in metabolism across the brain. These results show that with appropriate magnetic field shimming, the ^2^H-glucose and ^2^H-water peaks were easily observable in all subjects using both MRS and MRSI acquisitions, despite only 16 Hz of spectral separation between the two peaks at 3 T and relatively broad linewidths (12.1 ± 1.2 Hz). This work could not separate ^2^H-lactate from ^2^H-lipids, although the 1.35 ppm peak increased by 93% in the first experiment, suggesting that about half the measured peak is likely from the formation of lactate. This contrasts with a recent study in humans at 9.4 T (Ruhm et al., 2021), which found the lipid signal exceeded lactate, which may be the result of using a surface array coil for reception. In comparison with the birdcage coil used here, surface arrays greatly amplify signal from scalp lipids. In comparison, the rapidly increasing ^13^C-lactate signal over time could be easily distinguished on ^13^C-HPMRI without lipid contamination.

In addition, this study has compared DMI and ^13^C-HPMRI in humans for the first time. An important finding of this study is that a relative difference in oxidative and glycolytic metabolism was observed using the two techniques, with ^13^C-HPMRI probing early rapid lactate production, and DMI probing the later slower production of Glx. Intravenous administration of ^13^C-pyruvate enabled rapid metabolite uptake and high SNR. Hyperpolarized ^13^C-pyruvate is rapidly transported across the normal blood-brain barrier, and within the cell it is metabolized into both ^13^C-lactate and ^13^C-bicarbonate, with marked regional variations in metabolism observable across the brain (Dwork et al., 2021; Ehrhardt et al., 2021; Grist et al., 2019; Lee et al., 2020; Ma and Park, 2020). A previous study has compared DMI and ^13^C-HPMRI pre-clinically using a 4.7 T system (von Morze et al., 2021), but given that the ^13^C-bicarbonate signal was undetectable, a direct comparison of the glycolytic/oxidative metabolism using each method could not be undertaken. Our results have shown that DMI is weighted towards the detection of oxidative metabolism, based on the low (<1) ^2^H-lactate/^2^H-Glx ratio, whereas ^13^C-HPMRI reveals higher glycolytic metabolism based on its higher (>1) ^13^C-lactate/^13^C-bicarbonate ratio. These contrasting results can be explained by differences in administration, cerebral uptake, timing and length of acquisition, and cellular physiology of the two probes. For example, the long time between oral administration of ^2^H-glucose and imaging in DMI, and the relatively slow and continuous uptake from the gastrointestinal tract, results in a measurement of metabolism which is more likely to reflect steady-state conditions rather than an immediate response to short-term perturbation.

The results demonstrate that at least an hour is required to probe steady-state metabolism after oral ^2^H-glucose administration: the initial dynamic study over 1 hour revealed that ^2^H-Glx continued to rise even at the end of the first hour. **Figure S1** and **Table 1** show that for the cases where the time from administration to imaging was 100 mins or more, the ^2^H-lactate/^2^H-Glx ratio was calculated to be within a relatively narrow range of 0.10-0.18, suggesting that steady-state had been reached by this timepoint. This ratio was 0.25-0.44 for the two timepoints below 100 mins, suggesting that it takes longer for the labelled glucose to reach steady-state following entry into the TCA cycle, than to reach steady-state with cytosolic lactate. An extrapolation from these results is that oral administration of glucose may be preferable to intravenous glucose to assess steady-state metabolism. There is further support for this hypothesis from the finding in a previous preclinical DMI study (von Morze et al., 2021) where the ^2^H-glucose was administered intravenously, where the lactate-to-Glx ratio was more than two-fold higher (0.46) than the ratio observed here using oral glucose (0.18±0.09), although other differences between rodent and human brain metabolism may also contribute to this finding. In this study, we did not demonstrate a significant change in blood glucose before and after oral glucose administration of ^2^H-glucose, which also suggests that this approach is probing steady-state metabolism. Future work directly comparing DMI measurements using both oral and intravenous glucose administration will be important to address how the route of administration may affect metabolic measurements.

In contrast to DMI, ^13^C-HPMRI is performed rapidly in seconds to minutes following intravenous injection of a supraphysiological concentration bolus of ^13^C-pyruvate. Therefore, metabolism measured with ^13^C-HPMRI is very dependent on the ^13^C-pyruvate delivery to the brain with higher signal detected within the well-perfused gray matter in comparison to white matter (Grist et al., 2019). The formation of downstream metabolites is also limited by transport across the BBB and into astrocytes or neurons. Furthermore, supraphysiological blood and tissue concentrations of pyruvate may transiently alter endogenous levels of intracellular pyruvate or lactate and may therefore probe an acute perturbation of metabolism. Therefore, the kinetics of pyruvate metabolism are a function of pyruvate delivery, MCT expression, cytosolic LDH activity and tissue lactate concentration, all of which may show regional and cellular variation across the brain. In comparison, glucose metabolism is dependent on glucose delivery, GLUT transporter expression, as well as enzymatic activity and metabolite concentrations within the glycolytic pathway and the TCA cycle.

The tissue and cellular origins of the metabolites detected with both DMI and ^13^C-HPMRI is unknown and may also account for the differences detected with the two techniques, which is an interesting area for future research. Both glucose and pyruvate are initially transported from the plasma across the luminal and abluminal surfaces of the endothelium. GLUT1 is the major glucose transporter across the BBB with higher concentrations on the abluminal surface compared to the luminal surface which facilitates transport of glucose down its concentration gradient into the brain parenchyma (Duelli and Kuschinsky, 2001). Astrocytic end-feet have high concentrations of GLUT1 and cover a large proportion of the capillary walls to facilitate rapid glucose transport into the brain, while GLUT3 is the major glucose transporter on neurons with different kinetic properties (Maher and Simpson, 1994). Therefore, the downstream metabolic products of glucose metabolism detected with DMI may arise from within the endothelial cells of the BBB, the surrounding astrocytes, or may have passed through both cell types before undergoing metabolism by neurons, microglia, or other subpopulations. Similarly, the ^13^C-pyruvate used in ^13^C-HPMRI takes a related journey across the BBB, transported in and out of the endothelial cells, mediated predominately by the monocarboxylic acid transporter isoform MCT1 (encoded by the gene SLC16A1) which transports endogenous lactate and ketone bodies across the BBB (Gerhart et al., 1997). Astrocytes express MCT1 and MCT4, and neurons express MCT2 which all have different kinetic properties for pyruvate transport (Pellerin et al., 2007). An interesting biological complication in identifying which cell type may dominate the metabolic signal detected with both DMI and ^13^C-HPMRI is that lactate itself may be exported from astrocytes and used as a neuronal fuel source (Magistretti et al., 1999). This lactate in turn may be converted into pyruvate and subsequently glutamate within the neuron, with release of the latter into the synaptic cleft. Finally, excess lactate may be exported by MCTs into the cerebral venous sinuses, particularly if there is a large concentration gradient to facilitate this, and the distribution of the hyperpolarized ^13^C-HPMRI in the midline and occipital regions is suggestive of a significant contribution from the venous contribution (Grist et al., 2019). In summary, the cellular differences in uptake and metabolism between pyruvate and glucose may partly explain the differences between the two techniques.

The direct comparison of the ratio of glycolytic to oxidative metabolism using both DMI and ^13^C was enabled by the sensitive detection of ^2^H-Glx and ^13^C-bicarbonate using spectroscopic imaging techniques. We performed both ^2^H and ^13^C measurements using a dynamic spectroscopic imaging sequence similar to that used previously (von Morze et al., 2021). There were some significant differences in the human results presented here and the previous preclinical study: there was an inverse correlation between ^13^C-lactate and ^2^H-Glx preclinically but not clinically, and ^13^C-bicarbonate was detected in all healthy volunteers but was below the detection limit in the rat studies. The differences in bicarbonate production may be explained by the effect of anesthesia on the BBB and its depression of oxidative metabolism in animals, compared to the awake imaging undertaken in humans (Hyppönen et al., 2021; Josan et al., 2013), as well as differences between species in permeability of the BBB to pyruvate (Miller et al., 2018). Additionally, primates have significantly more white matter than rodents (Mota et al., 2019), and the increased number of astrocytes may be responsible for greater bicarbonate production (Choi et al., 2012).

Proton (^1^H) is the only MRI nucleus in routine clinical use because of the high SNR that can be achieved due to its abundance *in vivo* and large gyromagnetic ratio (γ_1H_=42.6 MHz/T). The latter enables high equilibrium spin polarization and therefore signal, whereas carbon-13 and deuterium both have lower gyromagnetic ratios and *in vivo* abundance (γ_13C_=10.7 MHz/T; γ_D_=6.5 MHz/T). DMI with oral glucose offers potential advantages including the lower cost of labelled substrate compared to intravenous ^13^C-labelled probes, ease of implementation, and the only additional hardware required is a dedicated coil tuned to the deuterium frequency. In comparison, ^13^C provides higher resolution and signal-to-noise, enabling rapid temporal resolution of the hyperpolarized metabolites due to the increase in SNR of more than four orders of magnitude (Ardenkjær-Larsen et al., 2003), and has a short acquisition time which can be more easily accommodated into the routine patient pathway. Deuterons cannot be usefully hyperpolarized due to their short T_1_ (<300 ms) compared to that for ^13^C (>300 ms), and therefore cannot achieve a comparably high signal-to-noise within the timescale required for tissue metabolism. The counterpoint for the high magnetization of hyperpolarized ^13^C is that it is irreversibly depleted after sampling, so while ^13^C-HPMRI allows for rapid assessment of dynamic metabolism, DMI can utilize multiple signal averages to enhance SNR over a longer timeframe. ^13^C-HPMRI also requires additional hyperpolarization equipment and pharmacy preparation, which limits the number of sites that can undertake this type of imaging. These two methods therefore have separate strengths for probing oxidation and glycolysis, and taken together, are highly complementary and provide a greater understanding of tissue metabolism.

Other approaches have been reported for imaging glucose uptake and/or metabolism with MRI include glucoCEST (De Feyter et al., 2018; Nasrallah et al., 2013; Walker-Samuel et al., 2013) and ^1^H-MRSI (Glunde et al., 2011). These do not require additional contrast agents or hardware but rely on spectral properties of protons which can be susceptible to field non-uniformities, T_1_-weighting, and magnetization transfer effects. The lower gyromagnetic ratios of ^2^H and ^13^C are less sensitive than ^1^H to magnetic field non-uniformities, and we applied ^2^H- and ^13^C-specific eddy-current correction prior to MRSI to improve our spectral resolution further (McLean et al., 2021). In addition, DMI and ^13^C-HPMRI detect exogenous labelled molecules against a low background signal, which improves spectroscopic specificity. Although glucoCEST involves administration of unlabelled glucose, the change above background signal is modest, and it cannot distinguish downstream metabolites as is the case with DMI, ^13^C-HPMRI, and ^1^H-MRSI. Despite the very high sensitivity of PET, the ^18^F-FDG glucose analog used for metabolic imaging is transported, phosphorylated, and trapped intracellularly, but does not undergo downstream metabolism and therefore does not provide direct information on glycolysis or oxidation. DMI and ^13^C-HPMRI present significant advantages over these techniques to image glucose metabolism in new ways.

The safety of injection and ingestion of the labelled probes is important for both DMI and ^13^C-HPMRI. Safety checks were performed prior to oral consumption or intravenous injection. Deuterium toxicology has been studied for nearly a century and is generally considered safe at the concentrations used in this study (Bachner et al., 1964): deuterium labelled compounds are used regularly in pharmaceutical compounds to improve drug safety through the strengthening of covalent bonds, which reduces the potential for forming toxic metabolites (Timmins, 2014). The quantity of glucose required for DMI is equivalent to that commonly used in the glucose challenge test for diabetes mellitus (Beysen et al., 2007), which is 15 g more than the maximum used in this study.

We have here demonstrated the practicability of DMI in human brain at 3 T, although there were several limitations in our implementation. Our quantification assumes a baseline concentration of deuterated water of 10.9 mM (Ashwal et al., 2004), and a similar approach has been followed in other studies (Veltien et al., 2021). However, baseline measurements before glucose ingestion were not routinely made here, confounding the estimation of absolute metabolite concentrations due to the conversion of ^2^H-glucose to HDO. The actual *in vivo* abundance of ^2^H may also vary due to atmospheric and environmental conditions (Merlivat and Jouzel, 1979). Absolute quantification and higher precision could be achieved using external calibration phantoms and correcting for regional coil sensitivity. However, the robustness of external calibration can be limited by variations in shim, temperature, B_1_ uniformity, and relaxation between the phantom and brain. Additionally, a calibration phantom included within the field of view might have been difficult to separate from the brain on imaging, due to the relatively coarse spatial localization used with an MRSI matrix of only 10×10×10. Because of this, we chose a ^13^C-MRSI sequence for comparison with a similar matrix of 10×10, albeit over a smaller FOV (20 cm for ^13^C-HPMRI vs. 32 cm for DMI). ^13^C-HPMRI demonstrated differences between gray and white matter which were not seen with DMI and may have been identified if a higher spatial resolution was used. One study at a higher field strength of 9.4 T demonstrated higher Glx continuing to increase in gray matter compared to white matter up to two hours after oral administration of ^2^H-glucose (Ruhm et al., 2021). The intravenous administration of ^2^H-glucose would result in higher *in vivo* concentrations thus increasing SNR, enabling higher resolution DMI at 3 T.

## Conclusion

This study compared normal human brain metabolism using two emerging MRI techniques for the first time. We have demonstrated the potential of DMI at 3 T to measure orally administered ^2^H-labelled glucose metabolism in the normal human brain, discriminating spectral peaks which could be used to measure both oxidative and glycolytic metabolism. This was performed at a clinical field strength and within a clinically feasible timeframe of 10 min. The comparison between DMI and ^13^C-HPMRI in the same normal volunteer cohort demonstrated that DMI was weighted towards detection of oxidative metabolism, which may reflect steady-state cerebral metabolism. In contrast, ^13^C-HPMRI detected higher levels of glycolytic metabolism due to early dynamic metabolism of the intravenously injected hyperpolarized ^13^C-labelled pyruvate into lactate. These contrasting results can be explained by differences in the administration of the respective probes, timing of imaging after administration, as well as biological differences in cerebral uptake and cellular metabolism between glucose and pyruvate. The results demonstrate these two metabolic imaging methods provide different yet complementary readouts of oxidative and glycolytic metabolism in the human brain.

## Data Availability

Data are available through the correspondence to the authors, subject to the following restrictions:
The data will not be shared prior to formal publication in a non-preprint journal.
The data will be shared subject to a formal agreement that provides co-authorship to the original authors and declares the scope of the new study
A formal project outline should be submitted to help frame this agreement
Identifying data will not be shared
The code used an available open source toolbox (AMARES), which is already available to the community.

## Acknowledgments

This study was funded by Cancer Research UK (CRUK; C19212/A27150; C19212/A16628), the Multiple Sclerosis Society (2015-35), the National Institute of Health Research (NIHR) Cambridge Biomedical Research Centre (BRC-1215-20014), CRUK Cambridge Centre, the Cambridge Experimental Cancer Medicine Centre, the Evelyn Trust, Addenbrooke’s Charitable Trust, and the NIHR/Wellcome Trust Cambridge Clinical Research Facility. JDK is supported by the European Union’s Horizon 2020 research and innovation programme (grant agreement no. 761214). The views expressed are those of the authors and not necessarily those of the NHS, the NIHR, the EU H2020 programme, or the Department of Health and Social Care.

We are also grateful to Elizabeth Latimer for assistance in this project and to Dr James Grist for his work on the ethical application.

## Supplementary Material

**Supplementary Figure S1.**
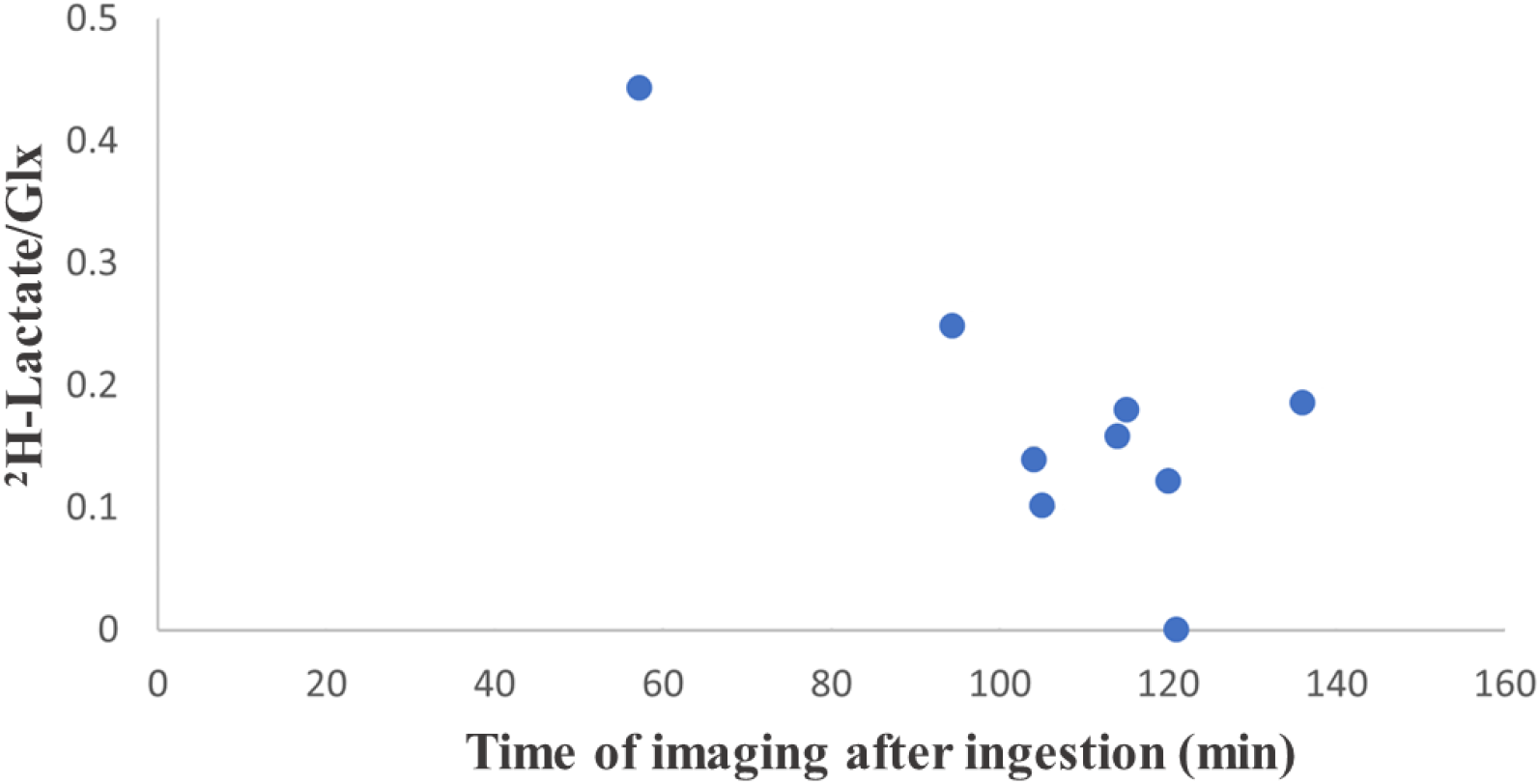
**Comparison of ^2^H-lactate/Glx on DMI compared to time after consumption of ^2^H-glucose. There is a linear correlation of ^2^H-lactate/Glx ratio and time, demonstrating that ^2^H-Glx production is slow and extends beyond an hour after ingestion (Pearson r^2^ = 0.69, p = 0.002).**

**Supplementary Figure S2.**
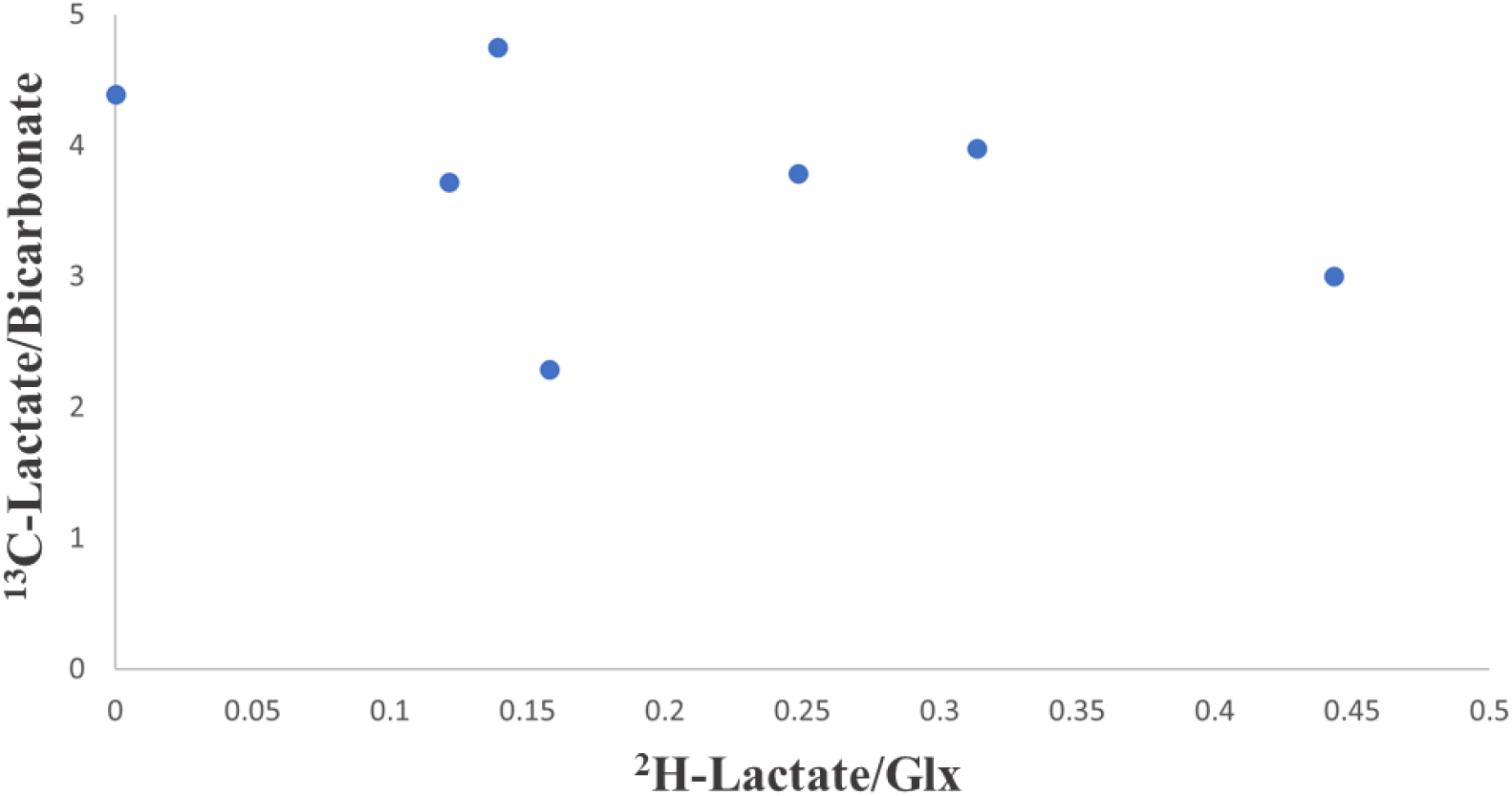
**A comparison of the ^13^C-lactate/bicarbonate ratio derived from ^13^C-HPMRI and the ^2^H-lactate/Glx ratio from DMI. The differences in scale of the two axes reflect that the ^13^C-HPMRI measures are >1 while the DMI measures are <1, demonstrating the differential way in which these techniques report on cerebral metabolism.**

